# Managing electromyogram contamination in scalp recordings: an approach identifying reliable beta and gamma EEG features of psychoses or other disorders

**DOI:** 10.1101/2021.11.18.21265963

**Authors:** Kenneth J. Pope, Trent W. Lewis, Sean P. Fitzgibbon, Azin S. Janani, Tyler S. Grummett, Patricia A.H. Williams, Malcolm Battersby, Tarun Bastiampillai, Emma M. Whitham, John O. Willoughby

## Abstract

**Objective:** In publications on the electroencephalographic (EEG) features of psychoses and other disorders, various methods are utilised to diminish electromyogram (EMG) contamination. The extent of residual EMG contamination using these methods has not been recognised. Here, we seek to emphasise the extent of residual EMG contamination of EEG.

**Methods:** We compared scalp electrical recordings after applying different EMG-pruning methods with recordings of EMG-free data from 6 fully-paralysed healthy subjects. We calculated the ratio of the power of pruned, normal scalp electrical recordings in the 6 subjects, to the power of unpruned recordings in the same subjects when paralysed. We produced “contamination graphs” for different pruning methods.

**Results:** EMG contamination exceeds EEG signals progressively more as frequencies exceed 25 Hz and with distance from the vertex. In contrast, Laplacian signals are spared in central scalp areas, even to 100 Hz.

**Conclusion:** Given probable EMG contamination of EEG in psychiatric and other studies, few findings on beta- or gamma-frequency power can be relied upon. Based on the effectiveness of current methods of EEG de-contamination, investigators should be able to re-analyse recorded data, re-evaluate conclusions from high frequency EEG data and be aware of limitations of the methods.

## 1. Introduction

1.1 The aetiology of the psychoses likely involves a complex combination of genetic, neurodevelopmental, neurodegenerative and environmental factors and there are reasons to propose that high-frequency electroencephalographic (EEG) rhythms might be informative. For example, in addition to the positive and negative symptoms of schizophrenia, other symptoms include cognitive decline, and impaired cognitive processes such as working memory, which are based on synchronised neural oscillations, particularly in the beta/gamma (25-40+ Hz) frequency ranges (McCutcheon et al., 2019, Uhlhaas and Singer, 2013). These rhythms are generated by phasic activity in GABAergic interneurons which input onto fast-firing pyramidal neurons, and thus produce high-frequency beta/gamma oscillations (Deutsch et al., 2010, Reynolds et al., 2004). Patients with schizophrenia have fewer pyramidal cell dendritic spines (as shown in auditory and dorsolateral prefrontal cortex (Gonzalez-Burgos et al., 2015) which impair excitatory input to pyramidal cells and, thereby, reduce pyramidal cell activation, including onto GABAergic interneurons. The consequence is reduced recurrent inhibitory regulation of pyramidal cells (Coyle et al., 2012, Rujescu et al., 2006). Also potentially relevant to a GABAergic abnormality in schizophrenia is clozapine’s unique mechanism of action: compared to other antipsychotics for treatment-resistant schizophrenia, clozapine may have specific GABAergic properties (Nair et al., 2020). Evidence for beta/gamma EEG dysfunction in schizophrenia also comes from studies showing reduced auditory steady state responses (Brenner et al., 2009, O’Donnell et al., 2013, Onitsuka et al., 2013), so that aberrant beta/gamma activity likely underlies both the cognitive and negative symptoms of schizophrenia (Lewis et al., 2012).

1.2 There has been extensive investigation of beta/gamma activity in the standard resting electroencephalogram (EEG) in psychoses, but recent reviews have been unable to identify consistent beta or gamma EEG findings (Lavoie et al., 2019, Newson and Thiagarajan, 2018, Reilly et al., 2018). However, the problem of electromyogram (EMG) contamination plagues resting EEG beta/gamma in normal scalp recordings and could explain the absence of robust findings in resting EEG studies. We have examined this question by comparing EEG recorded before (EMG-contaminated) and after total neuro-muscular paralysis (EMG-free) in healthy volunteers. EMG contamination exceeds EEG power 5-to 200-fold in the 50-100 Hz range over much of the scalp, relatively sparing only the central scalp area and frequencies below 20 Hz (Pope et al., 2009, Whitham et al., 2008, Whitham et al., 2007).

1.3 Whilst various approaches are used to reduce EMG contamination (EMG “pruning” methods) in psychiatric studies (Reilly et al., 2018), testing the effectiveness of various methods against EMG-free data, reveals that none of the methods fully eliminate EMG contamination of resting EEG (Dharmaprani et al., 2016, Fitzgibbon et al., 2016, Fitzgibbon et al., 2015, Fitzgibbon et al., 2013, Janani et al., 2020, Janani et al., 2018a, Janani et al., 2017, Janani et al., 2018b). Thus, inconsistencies or negative findings may be due to biological variability in EMG contamination at different scalp locations and at different frequencies in different patient groups. Even when methods such as Independent Component Analysis (ICA) are used, an inadequate number of electrodes sometimes prevents their reliable application (Janani et al., 2018a).

1.4 Calculated-Laplacian estimations of current source density (CSD) addresses EMG contamination for the central scalp (Fitzgibbon et al., 2015, Fitzgibbon et al., 2013) and are methods that until now have infrequently been applied to continuous EEG.

1.5 To assist investigators in determining which published findings are likely to be based on valid, EMG-uncontaminated data, in this paper we show the possible extent of EMG-contamination of continuous EEG using a range of available EMG-pruning methods, from the minimally-processed through to the best available method. Currently, the latter is Canonical Correlation Analysis (CCA) used with a Support Vector Machine (SVM) to classify components, hence “pruned-CCA-SVM” (Janani et al., 2020). In this paper, we compare spectral power of pruned EEG data to EMG-free data and present the estimated residual contribution of EMG contamination in ‘contamination graphs’. These estimates in healthy subjects should be helpful in evaluating previously published work in psychoses and are salutory indicators of the severity of the problem. They may provide motivation for researchers to re-analyse their recorded data or re-evaluate their conclusions on high frequency EEG data recorded away from the central scalp.

## 2. Methods

### 2.1 Subjects

We applied a range of EMG-pruning methods to scalp electrical recordings from six healthy subjects and calculated the extent to which spectral power of the processed scalp electrical recording exceeded EMG-free EEG from the same subjects when paralysed. This data has been utilised in papers on the EMG contamination problem published by our group over the last 13 years, initially with two and, ultimately, with six subjects (Fitzgibbon et al., 2016, Fitzgibbon et al., 2015, Fitzgibbon et al., 2013, Janani et al., 2020, Janani et al., 2018a, Janani et al., 2017, Pope et al., 2009, Whitham et al., 2008, Whitham et al., 2007).

### 2.2 Data

EEG was recorded using a 128-channel system (Compumedics Pty Ltd., Melbourne, Australia) while the subjects, with pre-inserted laryngeal-mask airways, were at rest in a recliner-chair. They undertook various tasks, including resting Eyes-Closed, before and again after total neuromuscular blockade with a curare-like drug (Whitham et al., 2008, Whitham et al., 2007). [Note: the subjects were semi-recumbent with their head resting on the recliner-chair, so the extent of EMG-contamination of electrodes close to cervical muscles would be less than expected in studies with the head un-supported.]

### 2.3 Data Processing

First, we applied the EEGLab detrend algorithm from the SIFT toolbox (Delorme et al., 2011), to remove low-frequency noise, and the CleanRawData algorithm to remove channels with a low correlation (r < 0.85) relative to the other channels. The resulting data is what we will refer to as “EMG-contaminated” data. We then applied several EMG-pruning methods:

#### 2.3.1 ICA (Delorme and Makeig, 2004)

This identifies common signals (independent components) contributing to subsets of electrodes. These components may correspond, for example, to EEG, eye-movements, blinks, electrocardiogram, or muscle contraction, and the contaminants are selectively discarded. Electrode recordings can then be calculated without contributions from contaminating components. We used ICA Infomax from EEGLab and also used spectral slope to assist in identifying neural components (Cardoso, 1997). Spectral slope refers to the shape of the power spectrum of each component and assists in classifying a component as brain or muscle origin. EMG has more sustained power between 7 and 75 Hz (Goncharova et al., 2003), so has a ‘flatter’ slope than EEG.

#### 2.3.2 CCA or CCA-SVM with spectral slope (Janani et al., 2020)

This method utilises CCA to obtain components and their spectral slopes (De Clercq et al., 2006) and uses SVM to identify brain or non-brain components, including electrical noise (Dharmaprani et al., 2016, Fitzgibbon et al., 2016, Janani et al., 2020, Janani et al., 2018a).

#### 2.3.3 Calculated Laplacian (Fitzgibbon et al., 2013, Kayser and Tenke, 2015)

The Laplacian transform provides a different measure of brain activity than EEG. It calculates the amount of current emerging perpendicularly from the brain surface, ie scalp current density. For electrodes where there is no underlying cranial or scalp muscle, Laplacians reveals brain activity with minimal contaminating signal from muscle.

### 2.4 Estimates of residual EMG contamination

We calculated ratios of processed (unparalysed) spectral power to the EMG-free (paralysed) spectral power for commonly used EEG bands: delta (1-4 Hz), theta (4-8 Hz), alpha (8-13 Hz), beta (13-30 Hz), gamma1 (30-50 Hz and gamma2 (50-100 Hz), although we excluded 48-52 and 98-100 Hz power due to line noise and its first harmonic. We display the results as ‘contamination graphs’ using the 10-20 montage display of 21 channels to conveniently show the extent of residual EMG contamination. However, calculations were always made using all valid 128-channel data. The Laplacian transforms were made on all raw and pruned-EEG data and are displayed in separate figures. For both EEG and Laplacians, we show contamination graphs for EMG-contaminated data, pruned-CCA-EEGLab, pruned-ICA-EEGLab, and pruned-CCA-SVM.

### 2.5 Review of recent studies in psychoses

Using the contamination graphs (Figs 1 and 3) as a guide, we assessed papers that described analyses of resting beta- and gamma-EEG in psychiatric populations published since the reviews of Newson and Thiagarajan (2018) and Reilly et al., (2018). We excluded studies using time-locked signal averaging, intra-cranial, magnetoencephalographic (MEG) or non-human recordings, or those where no beta/gamma claims were made.

**Figure 1.**
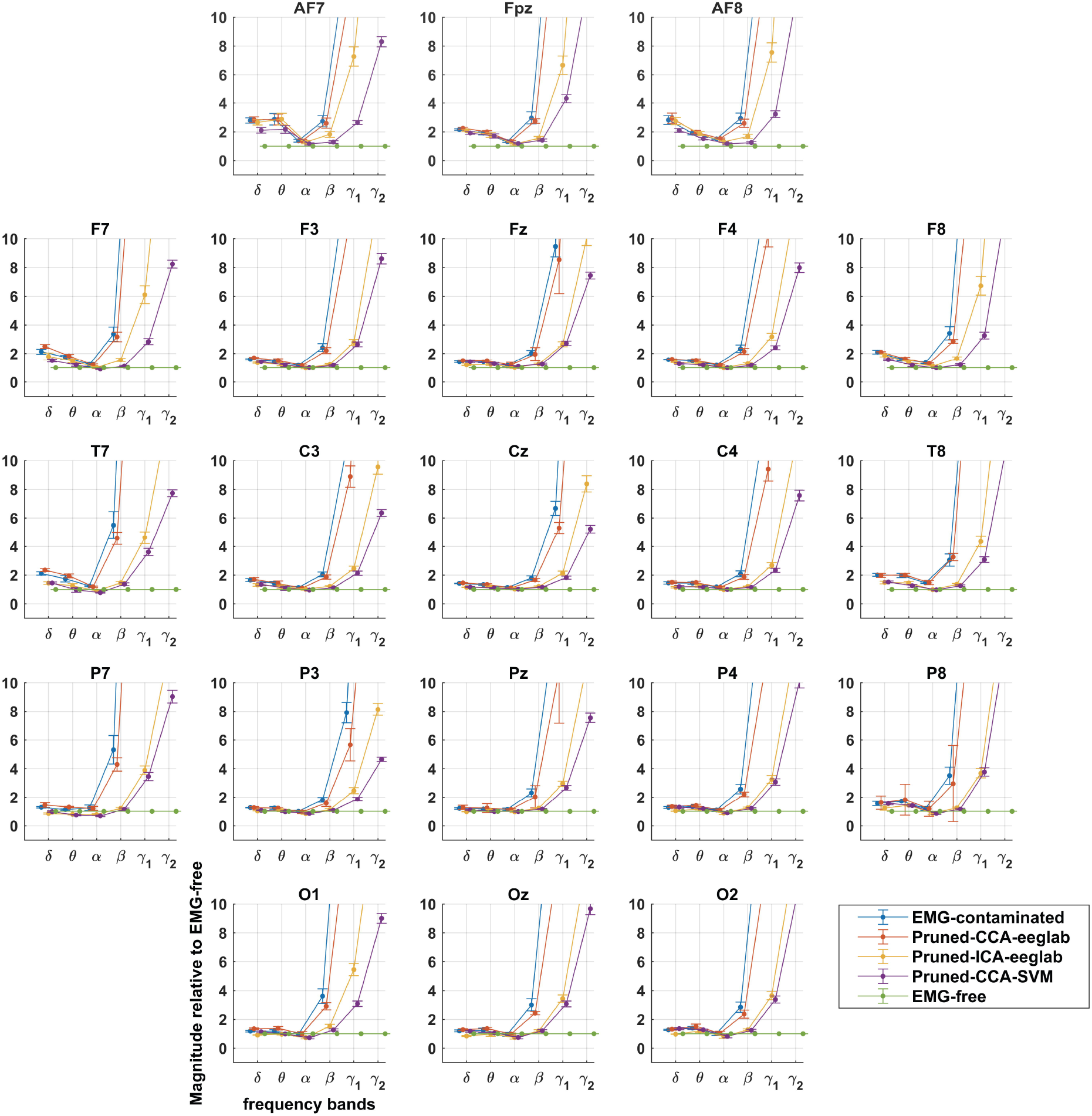
Montage showing extent of EMG-contamination of EEG relative to EMG-free recording with different pruning methods. Divergence from EMG-free is prominent in gamma bands and increased with increasing distance from the central scalp at Cz. Delta and theta bands show small increases, while alpha bands are reduced during paralysis (see also Fig. 3).

## 3. Results

### 3.1 Beta and Gamma

3.1.1 Contamination graphs for EMG-contaminated EEG (Fig.1) show EMG to be prominent in gamma bands (> 30 Hz) across the entire scalp. With Laplacian processing, however, there was limited or no EMG-contamination at central scalp sites. In the EEG, contamination also occurs to a lesser extent in the beta band, around 2-fold, and more if recordings are away from the central scalp (Figs 1 and 2). CCA-EEGLab-processed EEG remains contaminated almost as much as unprocessed EEG (Fig.1). For Laplacians, CCA-EEGLab provides a slight further improvement in contamination of the peripheral (EMG-contaminated) electrodes (Fig. 2).

**Figure 2.**
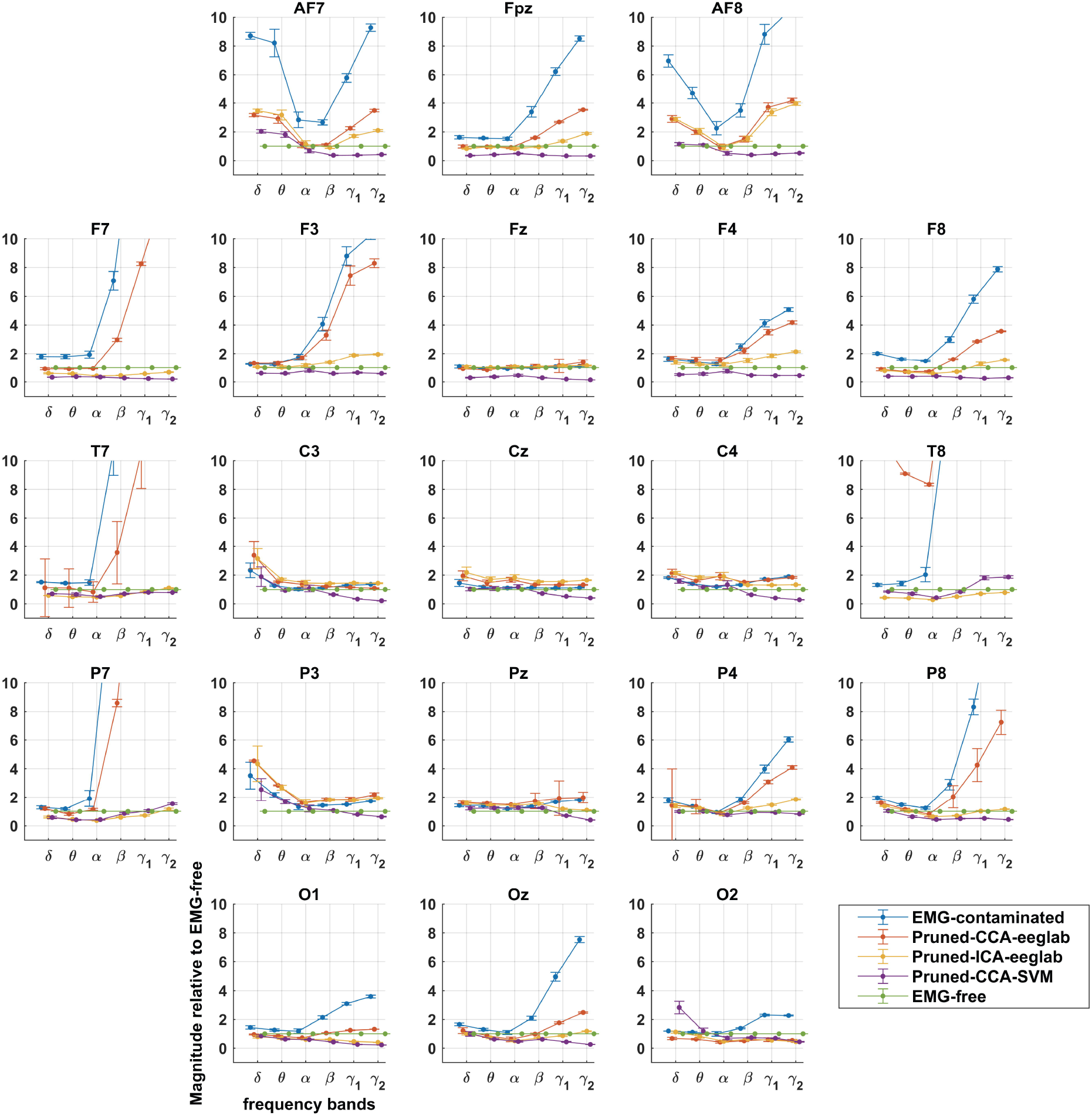
Montage showing extent of EMG-contamination of Laplacians relative to EMG-free recording with different pruning methods. Divergence from EMG-free is prominent in gamma bands and increased with increasing distance from the central scalp at Cz. Delta and theta bands also show increases, while alpha bands are reduced during paralysis (see also Fig. 4).

**Figure 3.**
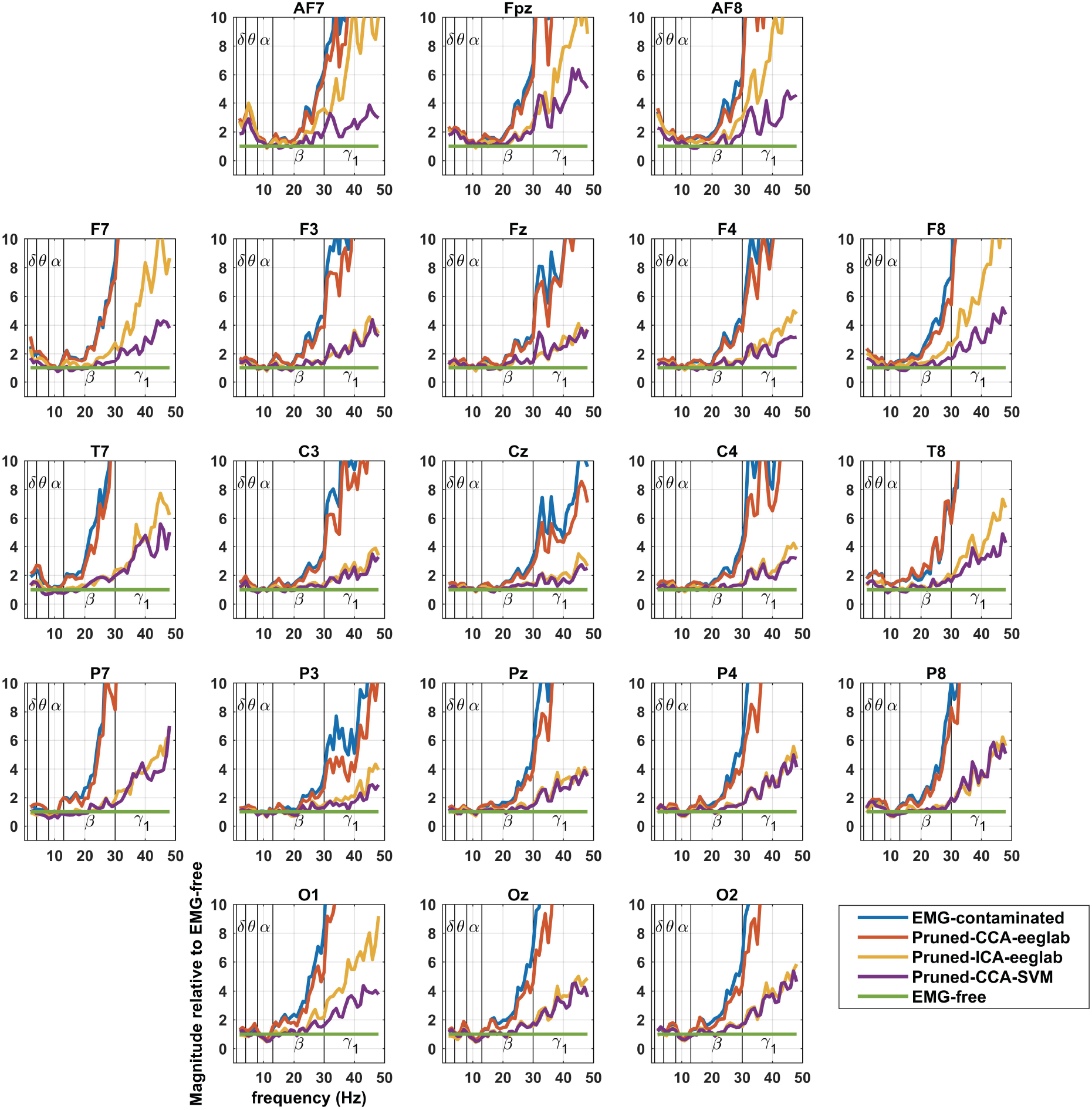
Montage showing extent of EMG-contamination of EEG relative to EMG-free to better display the frequency at which spectral power increases above EMG-free. Delta and theta frequencies also show increases, while alpha frequencies are reduced posteriorly during paralysis.

3.1.2 ICA-EEGLab achieves effective pruning of EMG in beta EEG activity in nearly all leads, however, gamma1 and 2 EEG remain heavily contaminated (Fig. 1). ICA-EEGLab sometimes markedly improves Laplacians in the few electrodes that are contaminated in the very lateral or posterior scalp (Fig. 2).

3.1.3. CCA-SVM achieves good pruning in beta EEG activity, even in lateral leads. Gamma EEG activity, however, remains severely contaminated (Fig. 1). In contrast, Laplacian transforms of CCA-SVM data almost eliminates EMG contamination over the entire scalp (Fig. 2). There is slightly less beta and gamma power for CCA-SVM than EMG-free, possibly related to reduced contamination by electrical line-noise identified by the SVM (see Discussion).

Thus, for EEG, no pruning-method achieves beta and gamma band power equivalent to EMG-free across the scalp. In a large central scalp area and, in contrast to EEG, Laplacian data have very low levels of EMG contamination across the full spectrum to 100 Hz.

### 3.2 Delta through Alpha

In delta, theta and alpha frequencies, contamination graphs (see spectra in Figs 3 and 4) reveal little variation relative to EMG-free power, but slight increases or reductions from EMG-free can be seen. The increases (delta and theta) have a similar distribution to the distribution of the most serious contamination due to muscle, pointing to ongoing subtle muscle contractions in the unparalysed subjects (see Discussion). Alpha reductions have a posterior distribution, unlike the distribution of severe EMG contamination, so that an alternative explanation likely applies, possibly related to different mental states between the resting-unparalysed and resting-paralysed recordings (see Discussion).

**Figure 4.**
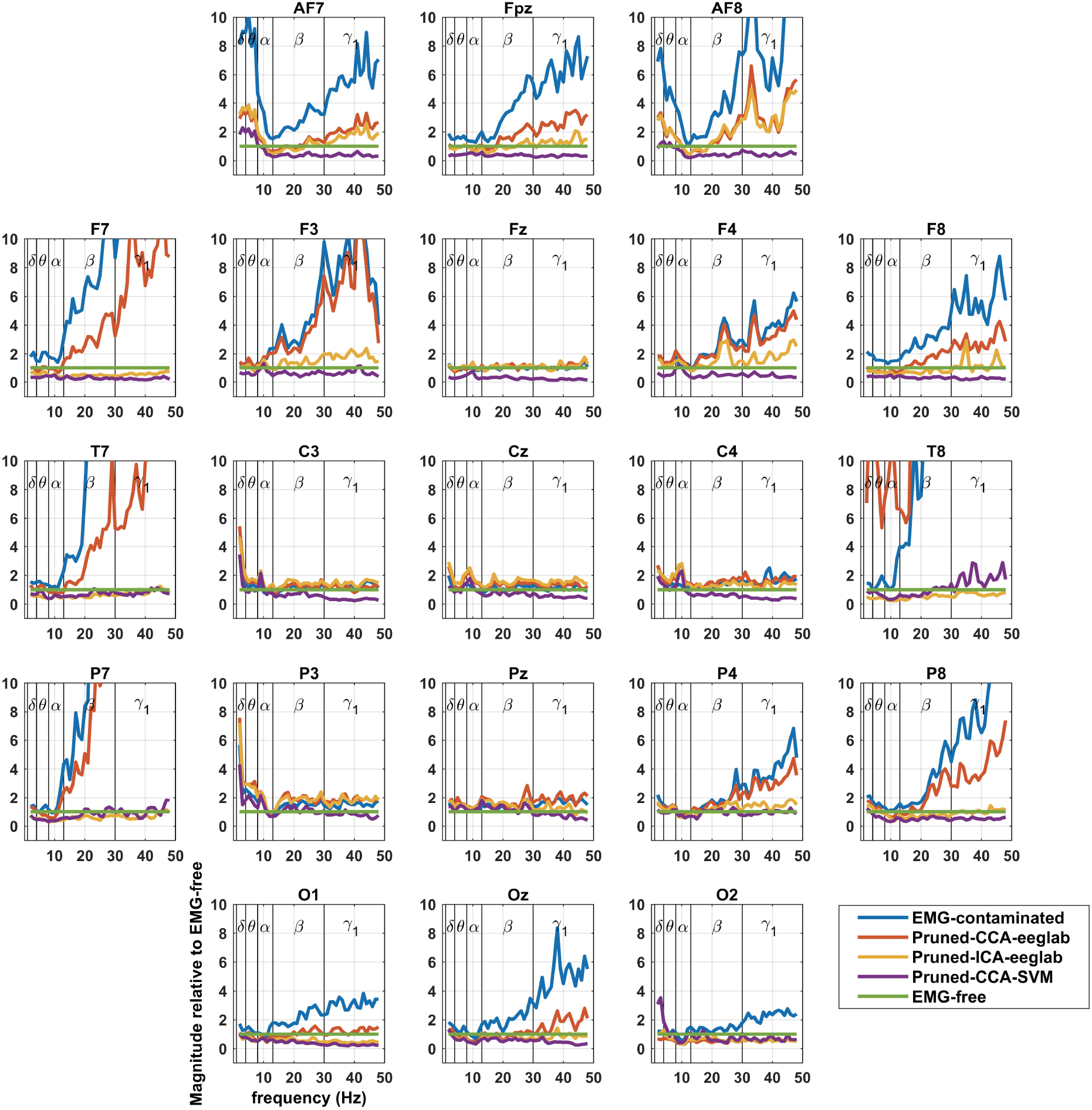
Montage showing extent of EMG-contamination of Laplacians relative to EMG-free to better display the frequency at which spectral power increases above EMG-free.

The frequencies at which divergence appears are around the beta/gamma boundary (25-35 Hz), displayed best in Figures 3 and 4. Again, the Laplacian results reveal minimal broad-spectrum contamination in central leads, and high levels of contamination very peripherally, except when pruning methods have been applied.

### 3.3 Pruning Methods

Of the methods we display here, Pruned-CCA-SVM performs best. Pruned-ICA-EEGLAB is next most effective, but diverges away from Pruned-CCA-SVM, most prominently where EMG activity is normally prominent, in very anterior and lateral leads (viz., F7, AF7, Fpz, AF8 and F8).

Although we have displayed 21 channel montages, contamination graphs from more peripheral locations exhibited even more EMG contamination than displayed here (data not shown).

### 3.4 Review of recent literature of beta/gamma EEG in psychoses

there were no publications whose results were exclusively drawn from adequately-pruned data from central scalp areas below 30 Hz.

## 4. Discussion

4.1 It is clear that EMG contamination continues to confound the analysis of high frequencies in the resting EEG, even when studies utilise careful artefact editing of recordings. The contamination graphs illustrate the difficulties that investigators will have in interpreting their own data, a situation we previously experienced (Fitzgibbon et al., 2004, Willoughby et al., 2003a, Willoughby et al., 2003b). While increases in gamma or beta EEG power could theoretically be measured above baseline EMG power, attributing a signal increase to a brain effect, rather than to a small change in the 5- to 40-fold baseline muscle power, is an insecure way to discover EEG correlates of psychoses or any other disorder.

4.2 Considering the EEG spectrum: the frequency at which power diverges from EMG-free EEG (best shown in Fig. 3) is approximately 20 Hz in unprocessed recordings and, after processing, approximately 40 Hz in much of the central scalp with both CCA-SVM and ICA-EEGLab. The useful range of EEG frequencies is roughly doubled by the use of these pruning methods. As previously published, the most effective of the pruning methods is CCA-SVM: it permits the interpretation of EEG spectra at slightly higher frequencies and in a slightly larger area than ICA-EEGLab (Janani et al., 2020). The slight reduction in beta and gamma relative power with CCA-SVM Laplacians, may be related to CCA-SVM identifying noise components in the line-voltage (Janani et al., 2020).

4.3 Considering Laplacian data: applying the Laplacian transform to derive current source density is clearly effective in avoiding EMG contamination in central scalp areas, though not peripherally when close to underlying muscles (Figs 2 and 4). At central sites, its effectiveness across the spectrum is a clear advantage over all other methods (compare Figs 3 and 4) and, furthermore, in the sub-50 Hz range, it extends the area of effectiveness to a larger central area than that provided by CCA-SVM EEG.

The advantage of the Laplacian method for central scalp sites provides motivation for analysis of Laplacian recordings as a routine in disease conditions. In this study, Laplacians were calculated from 120 channel EEG, covering the entire scalp, and enabled EMG-free measurement of brain activity in central scalp areas. Clearly, there would be a distinct advantage in recording Laplacians where this can be done reliably. The development of a tripolar concentric ring electrode, that records the Laplacian directly in hardware, offers this prospect (Koka and Besio, 2007) and is something we plan to evaluate.

### 4.4 Considering other bands

the small increases in relative power in delta and theta bands pre-paralysis, is consistent with delta and theta components being normal components of the resting EMG – as noted by Goncharova and colleagues (Goncharova et al., 2003). Alternatively, it is also possible that total paralysis causes its own ‘brain state’ characterised by reduced delta and theta activity. However, the low frequency increases in power occur with a similar distribution to the most serious contamination due to EMG (see Figs 1 and 3), consistent with EMG activity and/or with imperceptible motion artefact, for example, from other physiological activity such as respiration (Uriguen and Garcia-Zapirain, 2015). The small reductions in alpha power, relative to the paralysed state, have an occipital distribution and could possibly be attributed to enhanced mental alertness prior to being paralysed and then, after paralysis was fully established, increased relaxation, both physical and mental.

4.5 Since EMG contamination confounds studies of resting EEG, how can investigators safely proceed? We suggest general rules would be: start with at least 64-channel recordings; always prune resting EEG recordings, the best methods being Infomax EEGLAB and CCA-SVM (Janani et al., 2020); after pruning, apply Laplacian transforms to provide reliable data in central scalp areas. When confined to using EEG, if EEG power is increased at frequencies above 25 Hz, suspect EMG contamination, and if the finding becomes increasingly obvious away from the central cranium, assume the finding is EMG-confounded.

4.6 Currently the diagnosis of psychoses (schizophrenia, schizoaffective and bipolar disorder), is based only on clinical interview, collateral history and behavioural observation. There is no valid diagnostic test for psychoses like those that exist for other conditions (cancer, heart disease etc.) and delays in initial diagnosis impede efforts to initiate treatment early and reduce the duration of untreated psychosis. Reducing the duration of untreated psychosis can reduce long-term morbidity and improve quality of life. At present, the possibility of an abnormality in the resting EEG remains to be properly explored, specifically using Laplacian measurement. Such a finding would greatly assist diagnosis and likely further contribute to the current findings from The Bipolar and Schizophrenia Network for Intermediate Phenotypes (Tamminga et al., 2017) which is trying to develop a biomarker framework to categorise these conditions. It would further motivate drug discovery with a focus on GABAergic compounds for the treatment of psychoses: beta/gamma EEG abnormalities point specifically to GABAergic therapeutic interventions which might improve cognition and the negative symptoms of schizophrenia, which do not respond to current dopamine D2 antagonists. The question of beta/gamma resting EEG changes in psychoses requires further attention to aid in possible aetiological understanding, aiding earlier diagnosis, guiding prognosis and possibly monitoring response to pharmacological treatment.

## 5. Conclusion

Estimates of EMG contamination of scalp EEG in the range 1-100 Hz reveal that no method of EMG pruning adequately de-contaminates EEG data in the analysis of beta and gamma frequencies. Laplacian/current source density methods combined with pruning methods can provide EMG-free measures in central scalp areas. Based on the contamination graphs we have provided, it should be possible for EEG laboratories with signals-processing expertise to re-evaluate existing high-density EEG recordings using the methods we tested. For laboratories without signal-processing expertise, reliable conclusions should be possible using original data in safe frequencies and scalp locations.

## Data Availability

Graphical data produced in the present study are available upon reasonable request to the authors

## Acknowledgements

This work was supported by the National Health and Medical Research Council, the Flinders Medical Centre Foundation, the Clinician’s Special Purpose Fund of the Flinders Medical Centre, and an equipment grant from the Wellcome Trust, London, U.K.

